# Circulating extracellular vesicle-miRNAs as epigenetic mediators of metabolic, inflammatory, and neurological pathways in pediatric obesity

**DOI:** 10.1101/2025.07.16.25331410

**Authors:** Mangesh Dattu Hade, Eduardo Reátegui, J. Nicholas Brenton, Setty M. Magaña

## Abstract

Pediatric obesity represents a significant global health concern, associated with increased risk of early-onset metabolic disorders, chronic inflammation, and neurodevelopmental complications. Understanding the epigenetic mechanisms underlying obesity-induced metabolic and inflammatory dysregulation is essential for effective prevention and treatment. This pilot study provides the first comprehensive analysis of circulating extracellular vesicle (EV)-derived microRNAs (miRNAs) in pediatric obese individuals. Using age-and sex-matched pediatric obese (pOB) and healthy-weight controls (pNW), we identified distinct EV-miRNA signatures associated with pediatric obesity. Plasma EVs were isolated via tangential flow filtration and characterized using microfluidic resistive pulse sensing, transmission electron microscopy, and immunoblotting for canonical EV markers (CD81, CD63) and non-EV markers (calnexin). Differential miRNA profiling revealed significant obesity-driven epigenetic alterations, including known miRNAs involved in adipogenesis (miR-29a-3p, miR-643), lipid metabolism (miR-339-5p, miR-328-3p), inflammatory responses (miR-142-5p, miR-1249-3p), insulin signaling (let-7a-5p), and novel miRNAs (miR-1268a, miR-1268b, miR-3173-5p). Functional enrichment analyses using Kyoto Encyclopedia of Genes and Genomes and Gene Ontology highlighted significant regulation of pathways central to lipid metabolism, insulin resistance, mitochondrial function, immune activation, and neuronal signaling. Notable pathways, including PI3K-Akt signaling, MAPK signaling, calcium signaling, and axon guidance, were significantly enriched, emphasizing the systemic and central nervous system implications of the brain-fat axis in obesity. These findings underscore the potential of circulating EV-miRNAs as minimally invasive epigenetic biomarkers and epigenetic therapeutic targets for pediatric obesity. Larger studies are needed to validate the theranostic potential of EV-miRNAs in pediatric obesity.

## Introduction

Obesity is a present and escalating crisis around the globe, with 3.80 billion adults projected to be living with obesity by the year 2050[1]. The urgency of this global crisis is widely recognized; however, no country has yet succeeded in reducing adult obesity. One contributing factor to this increasing trend is the parallel rise in childhood and adolescent obesity[2]. Between 1990 and 2021, the combined prevalence of overweight and obesity among children and adolescents doubled globally, while obesity alone tripled[3]. Forecasts indicate that approximately 360 million children and adolescents aged 5–24 years will be living with obesity by 2050, a magnitude of burden that not only poses substantial immediate health risks but also significant economic implications, with total global economic impacts estimated to exceed 3% of the world’s gross domestic product by 2060[3].

The health consequences of pediatric obesity extend beyond metabolic dysregulation, profoundly influencing neurological functions. Disruptions of the blood-brain barrier (BBB) contribute to cognitive impairments and increased susceptibility to neurodegenerative conditions[4–6]. These findings underscore the importance of exploring the brain-fat axis, a bidirectional communication network connecting metabolic tissues with the central nervous system (CNS).

A key driver of obesity-related comorbidities is chronic low-grade inflammation resulting from dysregulated peripheral immunity. Impaired function of regulatory T cells (Tregs) contributes to metabolic dysfunction and sustained immune activation in adipose tissue[7,8]. The convergence of immune dysregulation, metabolic stress, and neurological consequences highlights the need for non-invasive biomarkers that capture the complexity of pediatric obesity.

Emerging areas of investigation in obesity-related chronic inflammation involve EVs, endogenous nanoparticles secreted by virtually all cell types, including adipocytes[9–11]. EVs are heterogeneous membrane-enclosed vesicles, such as exosomes and microvesicles, capable of exerting local and systemic effects by facilitating intercellular communication through their bioactive cargo (e.g., proteins, lipids, RNAs)[11,12]. Importantly, cargo loading into EVs is not random; rather, it is a tightly regulated and selective process reflecting the physiological or pathological status of the parent cell[13–15]. Thus, EV content can precisely mirror the functional state of tissues and serve as sensitive, minimally invasive biomarkers[15,16].

MicroRNAs , small non-coding RNAs that regulate gene expression post-transcriptionally, are increasingly recognized as critical epigenetic regulators of adipogenesis, lipid metabolism, immune signaling, and neuroinflammation[4,17]. Circulating within EVs, miRNAs are protected from degradation and capable of mediating intercellular communication, making them attractive candidates for liquid biopsy approaches. EV-miRNAs are particularly relevant due to their ability to reflect dynamic pathophysiological states and influence both peripheral and central systems[15,18].

Several miRNAs have been identified as direct regulators of epigenetic machinery or as targets of epigenetic modification themselves. For instance, miR-34a targets SIRT1, a histone deacetylase that governs chromatin remodeling and metabolic homeostasis; miR-29 family members regulate DNMT3A/B, modulating DNA methylation[19–22]. Additionally, miR-155 and miR-146a play roles in inflammatory feedback loops involving NF-κB and histone modifiers[23–25] , while let-7 miRNAs influence chromatin architecture through HMGA2, impacting adipocyte differentiation[26,27]. These examples highlight the dual role of miRNAs as both effectors and products of epigenetic regulation, underscoring their potential relevance to pediatric obesity.

While EV-miRNA alterations have been well-characterized in adult obesity and metabolic syndrome[28–32], studies in pediatric populations remain sparse. Adult studies have linked miRNAs, such as miR-122, miR-192, miR-223-3p, miR-33a, and miR-15b-5p, to metabolic signaling, inflammation, and therapeutic response[33–38]. However, pediatric obesity involves distinct developmental and immunometabolic features that warrant separate investigation.

These distinct developmental features become particularly critical given the forecasted acceleration in obesity prevalence among young children aged 5–14 years compared to older adolescents aged 15–24 years, indicating that earlier interventions targeting younger age groups might yield more significant long-term health benefits[1].

The brain-fat axis has emerged as a key conceptual framework for understanding how metabolic dysfunction can influence CNS health [39,40]. Epigenetic regulators like EV-miRNAs may play a central role in mediating this cross-talk, modulating pathways involved in appetite regulation, lipid homeostasis, inflammation, and BBB integrity[41]. By traversing biological barriers and influencing gene expression in both peripheral and central tissues, EV-miRNAs may be pivotal in linking early-life metabolic status with long-term neurodevelopmental outcomes.

In this pilot study, we present the first comprehensive profiling of circulating EV-derived miRNAs in pediatric obesity. We characterize distinct EV-miRNA signatures in obese versus healthy children and explore their potential regulatory roles in lipid metabolism, inflammation, immune homeostasis, and neuroimmune signaling. Emphasis is placed on the implications of these miRNAs for BBB integrity and cognitive function, reflecting the growing interest in the brain-fat axis. By integrating biomarker discovery with an epigenetic perspective, this study uncovers novel molecular pathways that may underlie pediatric obesity and its systemic and neurological consequences. Our findings offer new insights into how EV-miRNAs may mediate interorgan communication, shaping neurodevelopmental trajectories and informing the design of early, non-invasive diagnostic tools and precision therapeutic strategies tailored to the pediatric population. Given the rapid global increase of pediatric obesity highlighted by the latest forecasts, developing effective early diagnostic and interventional tools based on non-invasive biomarkers such as EV-miRNAs is particularly urgent [1].

## Results

### Enrolled study participants

For this pilot study, a total of six healthy adolescent subjects were included, categorized into healthy-weight (pNW, n = 3) and obese (pOB, n = 3) groups, matched by age and sex. Participants were aged between 17 and 20 years. The pNW group included two males and one female, all non-Hispanic, with an age range of 17–19 years and BMI ranging from 23.0 to 25.0 kg/m². The pOB group included one male and two females, all non-Hispanic, with an age range of 17–20 years and BMI ranging from 27.0 to 48.5 kg/m² (Table 1). All subjects had blood samples collected during early morning and while in the fasting state, with at least 8 hours of fasting prior to sample collection.

**Table 1:**
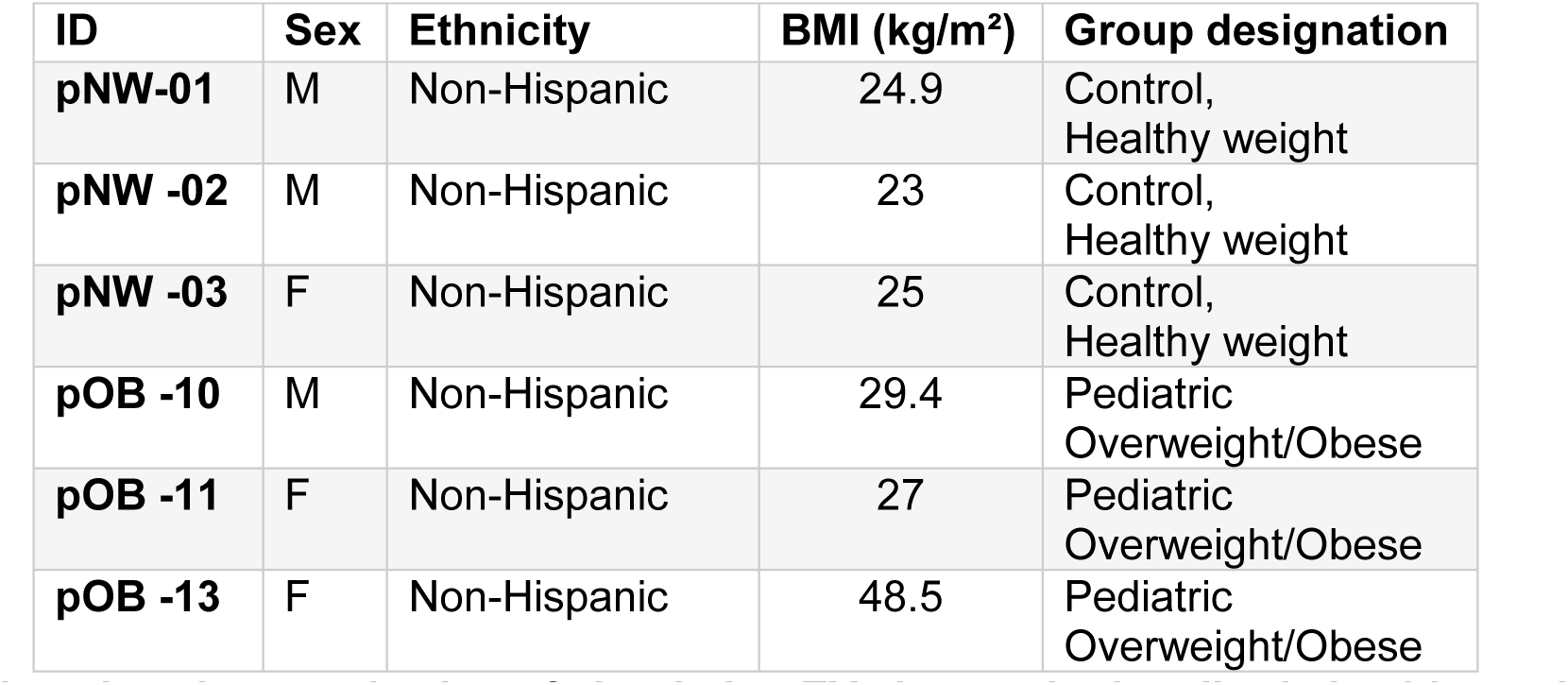
Participant demographics (pNW vs. pOB)

| ID | Sex | Ethnicity | BMI (kg/m<sup>2</sup>) | Group designation |
| --- | --- | --- | --- | --- |
| pNW-01 | M | Non-Hispanic | 24.9 | Control, Healthy weight |
| pNW -02 | M | Non-Hispanic | 23 | Control, Healthy weight |
| pNW -03 | F | Non-Hispanic | 25 | Control, Healthy weight |
| pOB -10 | M | Non-Hispanic | 29.4 | Pediatric Overweight/Obese |
| pOB -11 | F | Non-Hispanic | 27 | Pediatric Overweight/Obese |
| pOB -13 | F | Non-Hispanic | 48.5 | Pediatric Overweight/Obese |

### Comprehensive characterization of circulating EVs in matched pediatric healthy and obese individuals

We employed a standardized and optimized pipeline for the isolation and comprehensive characterization of circulating EVs from matched pediatric pNW and pOB individuals[42,43] (Figure 1A; see Materials and Methods). This methodology, adapted for pediatric plasma samples, integrates sequential centrifugation, thrombin-mediated defibrination, and tangential flow filtration (TFF) to ensure consistent and high-yield EV recovery.

**Figure 1.**
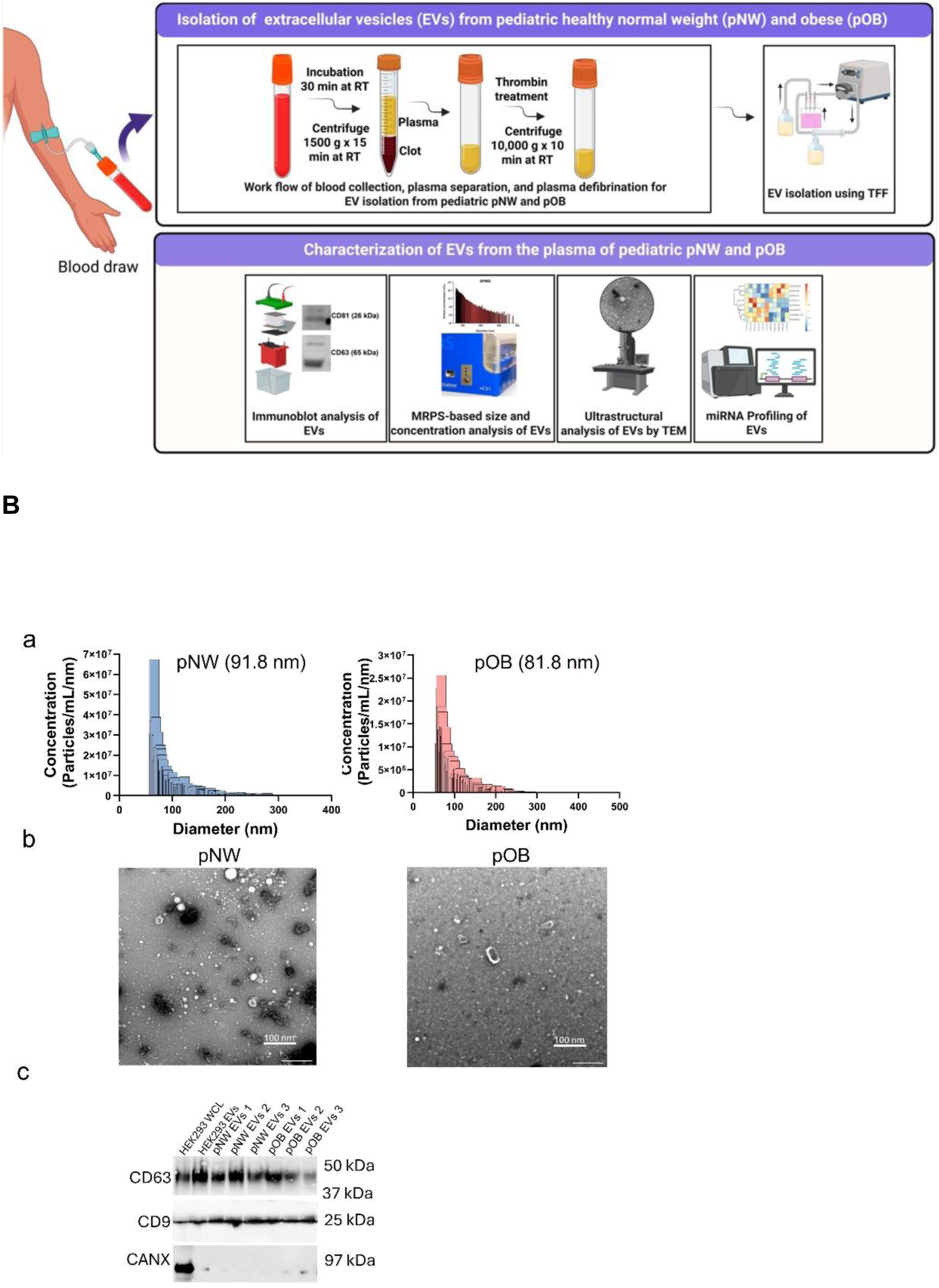
Isolation and multiparametric characterization of circulating EVs from matched pNW and pOB individuals. (**A**) EV isolation workflow. Whole blood was collected and incubated at room temperature (RT) for 30 min, followed by centrifugation at 1500 × g for 15 min to separate plasma. Plasma was defibrinated using thrombin and centrifuged again at 10,000 × g for 10 min at RT to remove residual fibrin. EVs were subsequently isolated from defibrinated plasma via TFF. Isolated EVs underwent multiparametric characterization by immunoblot for EV and non-EV markers, MRPS (particle sizing and concentration), TEM (ultrastructural morphology), and downstream miRNA profiling. (**B**) Detailed characterization of circulating EVs from pNW and pOB samples. (a) MRPS analysis showing comparable EV size distributions with median particle diameters of ∼71.7 nm (pNW) and ∼81.8 nm (pOB). Data represent three independent biological replicates per group. (b) Representative TEM images demonstrating heterogeneous populations of EVs exhibiting characteristic cup-shaped morphology in both groups (n = 3 independent biological replicates). Scale bar = 100 nm. (c) Immunoblot analysis confirming heterogeneous expression of canonical EV markers CD81 (26 kDa) and CD63 (65 kDa), and absence of the non-EV marker Calnexin (97 kDa) in plasma-derived EV samples from pediatric pNW and pOB groups (n = 3 independent biological replicates). pNW, pediatric healthy-weight; pOB, pediatric obese; TFF, tangential flow filtration; MRPS, microfluidic resistive pulse sensing; TEM, transmission electron microscopy.

To validate EV isolation and integrity, we conducted multiparametric EV characterization, including microfluidic resistive pulse sensing (MRPS), transmission electron microscopy (TEM), and immunoblotting. MRPS analysis revealed comparable EV size distributions in both pNW and pOB groups, with median particle diameters of approximately 71.7 nm for pNW and 81.8 nm for pOB individuals **(Figure 1Ba).** Particle concentrations demonstrated consistency across all biological replicates, confirming reproducibility and robustness of the isolation procedure.

Ultrastructural analysis by TEM consistently revealed heterogeneous populations of vesicles with characteristic “cup-shaped” morphologies across samples from both pNW and pOB pediatric groups **(Figure 1Bb).** Furthermore, immunoblot analysis confirmed the expression of canonical EV-associated markers, CD81 (26 kDa) and CD63 (65 kDa), concurrent with the absence of the non-EV marker calnexin (97 kDa) **(Figure 1Bc).**

Together, these results highlight the successful isolation and biophysical characterization of plasma-derived EVs from pediatric cohorts, establishing a reliable foundation for downstream molecular profiling and biomarker discovery in pediatric obesity.

### Obesity-driven alterations in EV-miRNAs reveal metabolic and inflammatory dysregulation

To investigate obesity-specific EV-miRNA repertoires, we performed differential expression analysis comparing pOB and pNW individuals. A total of 556 miRNAs were identified, of which 429 were shared between both groups. Additionally, 55 miRNAs were uniquely detected in pOB, and 72 miRNAs were exclusive to pNW (Figure 2A). Several miRNAs showed significant dysregulation, notably miR-5010-3p_R+1, miR-1249-3p, miR-328-3p, miR-339-5p, miR-197-3p, miR-296-5p, miR-15a-3p_1ss22AT, miR-1268b_R-2, and miR-1268a (upregulated), as well as miR-643, miR-29a-3p, miR-29c-3p, miR-142-5p_L+2R-2, and miR-27a-3p_R-1 (downregulated). Given the exploratory nature of this study and the small sample size, we applied a less stringent statistical threshold (p < 0.1) to define significant EV-miRNAs (**Supplementary Table 1**), while also evaluating EV-miRNAs at more conventional thresholds of p < 0.001, p < 0.01, and p < 0.05 (**Table 2**).

**Figure 2.**
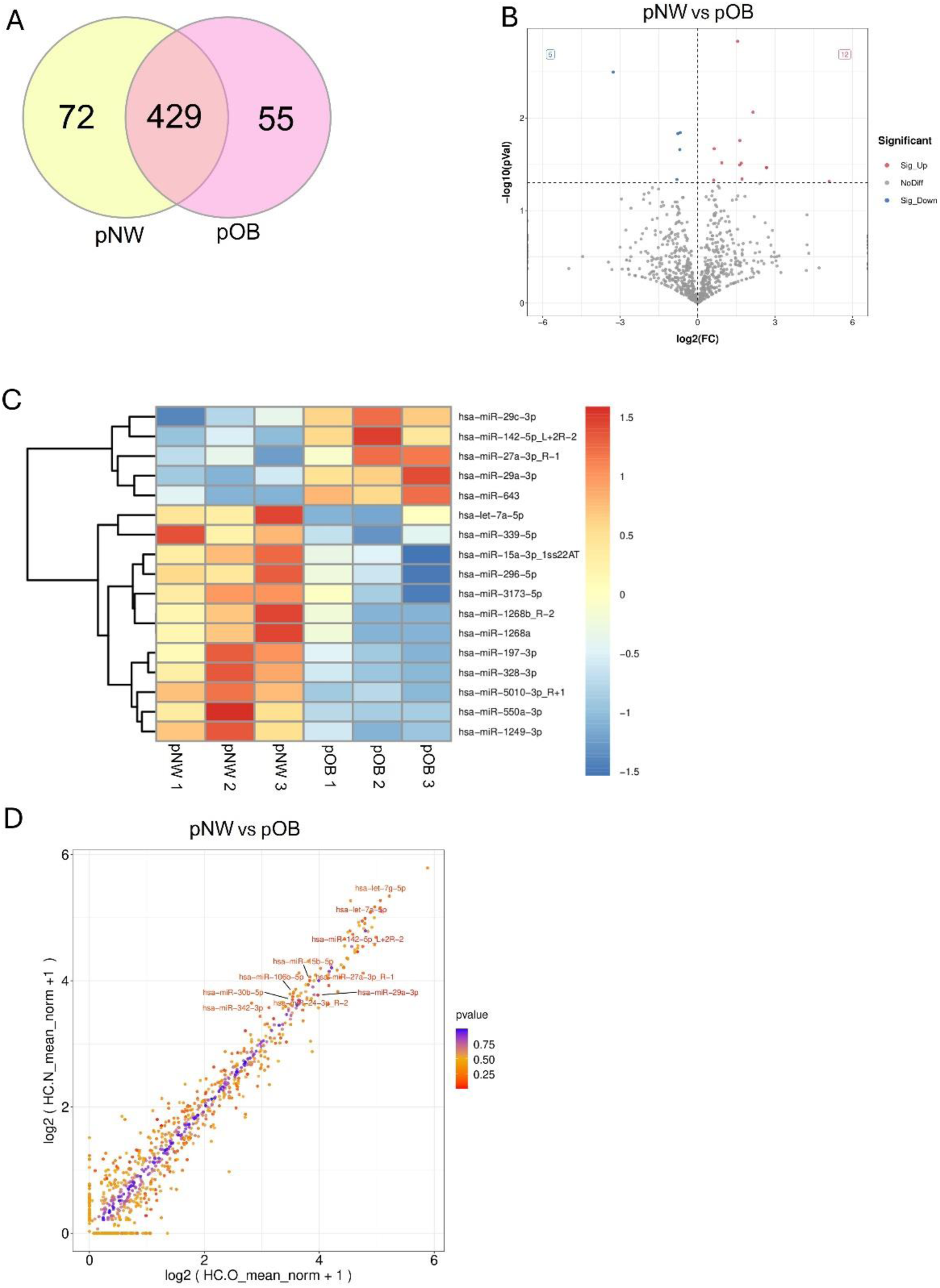
Global profiling of plasma-derived EV-miRNAs reveals distinct signatures in obese pediatric individuals compared to matched healthy-weight controls. (**A**) Venn diagram illustrating shared and unique EV-miRNAs in pOB and pNW. Of 556 identified miRNAs, 429 were shared, 55 were exclusive to pOB, and 72 exclusive to pNW. (**B**) Volcano plot depicting differential expression of EV-miRNAs between pOB and pNW. Significantly upregulated miRNAs in pOB are highlighted in red, while significantly downregulated miRNAs are shown in blue (cutoff criteria: |log₂ fold change| ≥ 1.2 and p ≤ 0.05). (**C**) Heatmap demonstrating hierarchical clustering of the top differentially expressed EV-miRNAs across individual pOB and pNW samples. Clustering was conducted using Euclidean distance. Expression levels are indicated in red (high expression) and blue (low expression), relative to median expression levels. (**D**) Scatter plot correlation analysis of EV-miRNA expression levels between pOB and pNW samples. Most miRNAs exhibit similar expression patterns across groups; however, certain miRNAs (e.g., miR-29a-3p, miR-142-5p, and let-7a-5p) show marked elevations specifically in the pOB group.

**Table 2.** Top differentially expressed EV-miRNAs in pNW vs. pOB

| Downregulated DEMs | FC | log2(FC) | P-value |
| --- | --- | --- | --- |
| miR-643 | 0.1 | -3.27 | 3.20E-03 |
| miR-29a-3p | 0.63 | -0.67 | 1.44E-02 |
| miR-29c-3p | 0.59 | -0.76 | 1.47E-02 |
| miR-142-5p_L+2R-2 | 0.62 | -0.69 | 2.20E-02 |
| miR-27a-3p_R-1 | 0.57 | -0.8 | 4.62E-02 |
| **Upregulated DEMs** |  |  |  |
| miR-550a-3p | 34.2 | 5.1 | 4.86E-02 |
| miR-1268a | 6.36 | 2.67 | 3.43E-02 |
| miR-1268b_R-2 | 6.36 | 2.67 | 3.43E-02 |
| miR-1249-3p | 4.43 | 2.15 | 8.67E-03 |
| miR-3173-5p | 3.29 | 1.72 | 4.56E-02 |
| miR-296-5p | 3.23 | 1.69 | 3.08E-02 |
| miR-328-3p | 3.11 | 1.64 | 1.76E-02 |
| miR-15a-3p_1ss22AT | 3.1 | 1.63 | 3.22E-02 |
| miR-5010-3p_R+1 | 2.93 | 1.55 | 1.49E-03 |
| miR-197-3p | 1.91 | 0.93 | 3.05E-02 |
| miR-339-5p | 1.56 | 0.64 | 2.15E-02 |
| let-7a-5p | 1.54 | 0.63 | 4.69E-02 |

DEM, Differentially expressed miRNAs; FC, fold change; pNW, pediatric healthy-weight; pOB, pediatric obese

Volcano plot analysis (Figure 2B) provides a global view of EV-miRNAs significantly upregulated (red) or downregulated (blue) in pOB compared to pNW (cutoff criteria: |log₂ fold change| ≥ 1.2 and p ≤ 0.05). Labeled EV-miRNAs represent significantly differentially expressed candidates. Hierarchical clustering of differentially expressed EV-miRNAs revealed clearly distinct heatmap expression profiles that differentiated pOB from pNW, suggesting obesity-specific EV-miRNA candidates (Figure 2C). Notable dysregulated miRNAs are listed in **Table 2**. Correlation analysis revealed similar expression patterns for most miRNAs across groups, while selected miRNAs (e.g., miR-29a-3p, miR-142-5p, and let-7a-5p) exhibited significant elevations, indicating potentially relevant miRNAs associated with obesity-driven metabolic dysregulation (Figure 2D).

### Functional enrichment analysis of differentially expressed EV-miRNAs in pediatric obese versus healthy-weight individuals reveals significant enrichment of metabolic, immune, and neuroregulatory signaling pathways

To identify biological pathways associated with altered EV-miRNAs in pOB compared to matched pNW, we performed KEGG pathway enrichment analysis. Significantly enriched pathways included pathways in cancer (P = 8.80 × 10⁻¹⁷, Q = 2.97 × 10⁻¹⁴, Rich Factor = 0.6912), ErbB signaling (P = 8.88 × 10⁻¹³, Q = 1.49 × 10⁻¹⁰, Rich Factor = 0.8837), Rap1 signaling (P = 2.05 × 10⁻¹², Q = 2.30 × 10⁻¹⁰, Rich Factor = 0.7535), axon guidance (P = 2.87 × 10⁻¹², Q = 2.42 × 10⁻¹⁰, Rich Factor = 0.7688), proteoglycans in cancer (P = 6.98 × 10⁻¹², Q = 4.70 × 10⁻¹⁰, Rich Factor = 0.7455), metabolic pathways (P = 9.95 × 10⁻¹², Q = 5.58 × 10⁻¹⁰, Rich Factor = 0.7337), focal adhesion (P = 1.27 × 10⁻¹¹, Q = 6.09 × 10⁻¹⁰), regulation of actin cytoskeleton (P = 1.47 × 10⁻¹⁰, Q = 6.19 × 10⁻⁹), calcium signaling (P = 3.84 × 10⁻¹⁰, Q = 1.43 × 10⁻⁸), PI3K-Akt signaling (P = 1.86 × 10⁻⁷, Q = 2.64 × 10⁻⁶), and MAPK signaling (P = 2.86 × 10⁻⁷, Q = 3.86 × 10⁻⁶) (Figure 3A**–C**).

**Figure 3.**
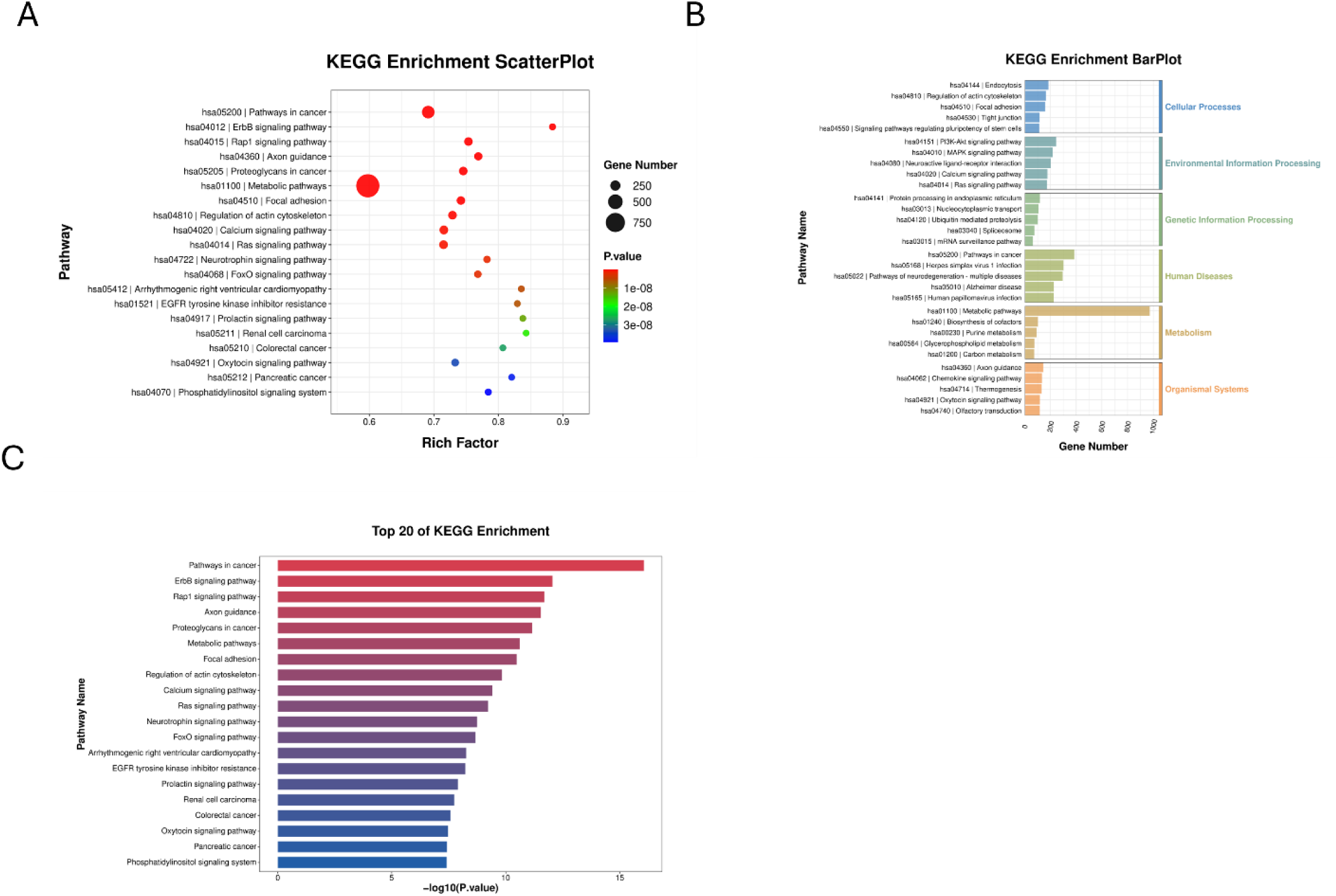
KEGG pathway enrichment analysis of differentially expressed EV-miRNAs in pediatric obese individuals versus those of healthy-weight reveals alterations in metabolic, inflammatory, and neuroregulatory signaling. (**A**) KEGG enrichment scatter plot illustrating significantly enriched pathways targeted by differentially expressed EV-miRNAs. Circle size corresponds to gene count per pathway, and color gradient represents adjusted P-value significance. Prominently enriched pathways include ErbB signaling, Rap1 signaling, axon guidance, calcium signaling, and regulation of actin cytoskeleton. (**B**) KEGG enrichment bar plot categorizing enriched pathways into major biological functions, including metabolism, environmental information processing, cellular processes, and organismal systems. Highlighted pathways include PI3K-Akt signaling, MAPK signaling, and focal adhesion. (**C**) Bar graph depicting the top 20 KEGG pathways ranked by significance (–log₁₀(P-value)). The most significantly enriched pathways include ErbB signaling, Rap1 signaling, axon guidance, calcium signaling, regulation of actin cytoskeleton, and MAPK signaling.

The KEGG enrichment scatter plot illustrates pathways with the highest enrichment signals based on gene count and statistical significance (Figure 3A). Categorization by functional domains (Figure 3B) highlighted enrichment across metabolism (e.g., metabolic pathways, cholesterol metabolism), cellular processes (e.g., focal adhesion, endocytosis), environmental information processing (e.g., PI3K-Akt, MAPK signaling), and organismal systems (e.g., oxytocin signaling, thermogenesis). The top 20 significantly enriched KEGG pathways ranked by –log₁₀(P-value) are shown in Figure 3C.

### Functional enrichment analysis of differentially expressed miRNAs in obese versus non-obese healthy pediatric individuals identifies disruptions in neuronal signaling, lipid metabolism, and mitochondrial functions

To identify biological processes, cellular components, and molecular functions associated with EV-miRNA alterations in pOB compared to matched pNW, GO enrichment analyses were conducted. GO enrichment scatter plot analysis (Figure 4A) revealed significant enrichment for terms including protein binding (Rich Factor = 0.5962, P = 0.00 × 10⁰⁰, Q = 0.00 × 10⁰⁰), cytoplasm (Rich Factor = 0.6332, P = 3.40 × 10⁻²⁸⁷, Q = 2.91 × 10⁻²⁸³), nucleus (Rich Factor = 0.6035, P = 5.57 × 10⁻¹⁸⁴, Q = 3.17 × 10⁻¹⁸⁰), cytosol (Rich Factor = 0.6217, P = 5.89 × 10⁻¹⁷⁵, Q = 2.51 × 10⁻¹⁷¹), nucleoplasm (Rich Factor = 0.6278, P = 4.38 × 10⁻¹²⁸, Q = 1.49 × 10⁻¹²⁴), metal ion binding (Rich Factor = 0.6253, P = 2.23 × 10⁻¹²¹, Q = 6.36 × 10⁻¹¹⁸), transferase activity (Rich Factor = 0.6754, P = 5.12 × 10⁻⁹², Q = 1.25 × 10⁻⁸⁸), membrane (Rich Factor = 0.5376, P = 2.91 × 10⁻⁸⁹, Q = 6.20 × 10⁻⁸⁶), nucleotide binding (Rich Factor = 0.6510, P = 3.88 × 10⁻⁷³, Q = 7.36 × 10⁻⁷⁰), ATP binding (Rich Factor = 0.6564, P = 2.63 × 10⁻⁶², Q = 4.49 × 10⁻⁵⁹), positive regulation of transcription by RNA polymerase II (Rich Factor = 0.6693, P = 1.85 × 10⁻⁵², Q = 2.11 × 10⁻⁴⁹), phosphorylation (Rich Factor = 0.7205, P = 7.26 × 10⁻⁴⁸, Q = 7.75 × 10⁻⁴⁵), kinase activity (Rich Factor = 0.7188, P = 3.60 × 10⁻⁴⁷, Q = 3.61 × 10⁻⁴⁴), DNA binding (Rich Factor = 0.5970, P = 2.86 × 10⁻⁴⁶, Q = 2.57 × 10⁻⁴³), and regulation of transcription by RNA polymerase II (Rich Factor = 0.6201, P = 1.01 × 10⁻⁴⁵, Q = 8.62 × 10⁻⁴³).

**Figure 4:**
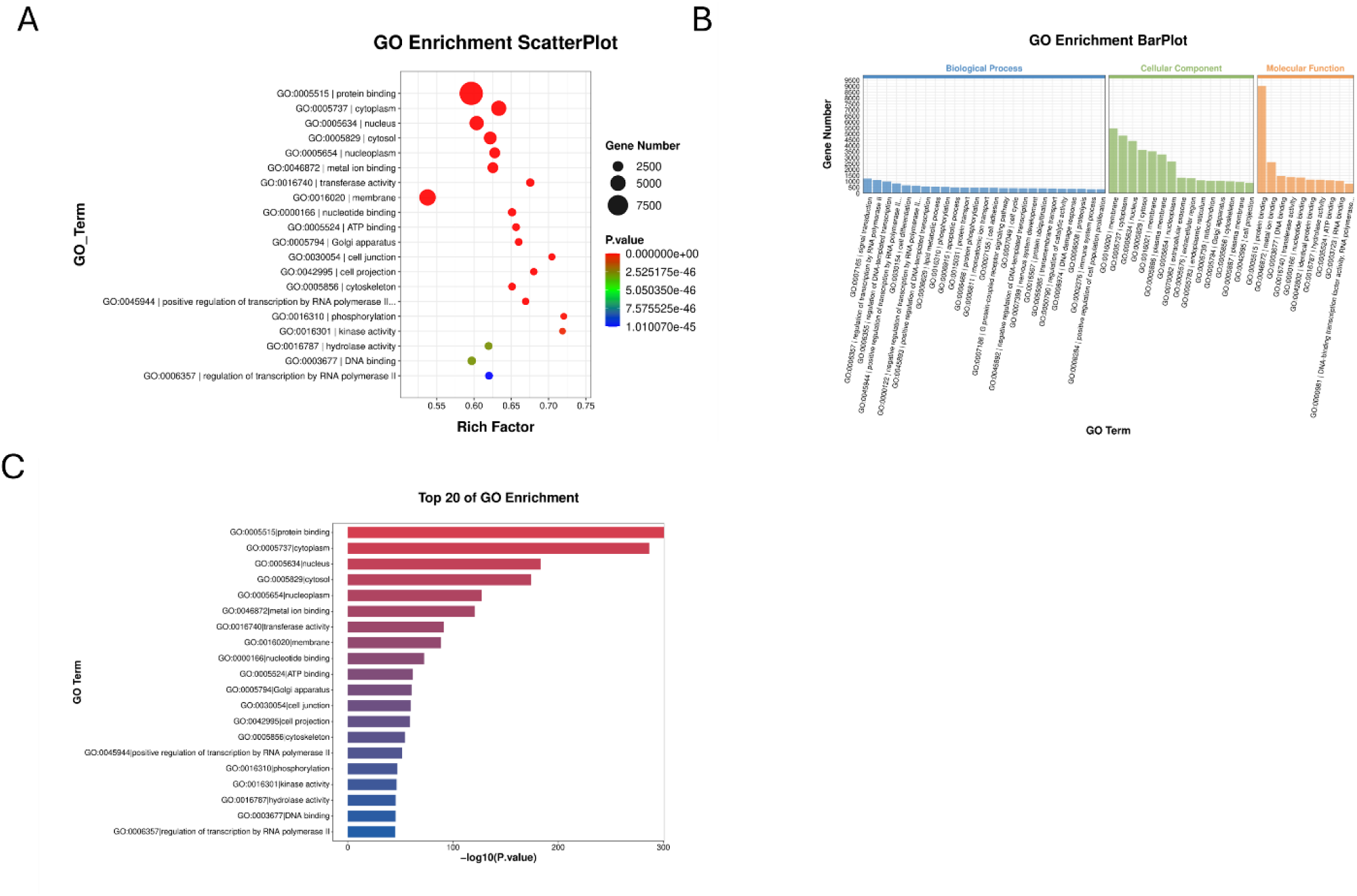
GO enrichment analysis of EV-miRNA targets in obese versus healthy-weight individuals reveals key disruptions in lipid metabolism, synaptic signaling, and intracellular regulation. **(A)** GO enrichment scatter plot illustrating significantly enriched Gene Ontology (GO) terms targeted by differentially expressed EV-miRNAs. Circle size corresponds to the number of genes associated with each GO term; color gradient represents the significance (adjusted P-value).(**B**) GO enrichment bar plot categorizing significantly enriched GO terms into three primary ontologies: Biological Processes, Cellular Components, and Molecular Functions.(**C**) Top 20 enriched GO terms ranked by significance (–log₁₀(P-value)), highlighting the most significantly enriched terms across all GO categories.

The GO enrichment bar plot (Figure 4B) categorized significantly enriched terms into biological processes, cellular components, and molecular functions. Biological processes included positive regulation of transcription by RNA polymerase II (Rich Factor = 0.6693, P = 1.85 × 10⁻⁵², Q = 2.11 × 10⁻⁴⁹), phosphorylation (Rich Factor = 0.7205, P = 7.26 × 10⁻⁴⁸, Q = 7.75 × 10⁻⁴⁵), protein phosphorylation, nervous system development, neuron projection, lipid metabolic processes, intracellular signal transduction, cell differentiation, and cell adhesion. Cellular components included cytoplasm (Rich Factor = 0.6332, P = 3.40 × 10⁻²⁸⁷, Q = 2.91 × 10⁻²⁸³), nucleus (Rich Factor = 0.6035, P = 5.57 × 10⁻¹⁸⁴, Q = 3.17 × 10⁻¹⁸⁰), cytosol, nucleoplasm, mitochondrion, endoplasmic reticulum, plasma membrane, and intracellular organelles. Molecular functions included protein binding (Rich Factor = 0.5962, P = 0.00 × 10⁰⁰, Q = 0.00 × 10⁰⁰), metal ion binding, transferase activity, nucleotide binding, ATP binding, kinase activity, and DNA binding.

The top 20 enriched GO terms ranked by significance (–log₁₀(P-value)) (Figure 4C) included protein binding, cytoplasm, nucleus, cytosol, nucleoplasm, metal ion binding, transferase activity, nucleotide binding, ATP binding, cell junction, cell projection, cytoskeleton, positive regulation of transcription by RNA polymerase II, phosphorylation, kinase activity, and DNA binding.

## Discussion

This pilot study provides the first comprehensive characterization of the circulating EV-miRNome in pediatric obesity. Our results revealed distinct EV-miRNA profiles implicated in obesity-driven metabolic, immune, and neuroimmune alterations, which may be unified by the brain-fat axis framework.

We observed marked dysregulation of key miRNAs known to epigenetically regulate adipogenesis, insulin sensitivity, and metabolic pathways. Specifically, miR-29a-3p, which modulates adipocyte differentiation and glucose homeostasis through DNA methyltransferases DNMT3A/B [20–22] [44,45], was notably dysregulated. The let-7a-5p miRNA family, associated with chromatin remodeling proteins such as HMGA2, was also significantly altered, impacting insulin signaling and metabolic regulation[26,27,46,47]. Additionally, the less characterized miRNAs, miR-643 and miR-5010-3p, exhibited significant differential expression, suggesting they may be potential new therapeutic targets for future research.

KEGG and GO pathway analyses revealed significant enrichment in critical metabolic signaling pathways, particularly the PI3K-Akt and MAPK signaling pathways, known for their roles in insulin sensitivity, glucose uptake, and energy metabolism[48,49]. Furthermore, enrichment of mitochondrial functions and intracellular organelle regulation pathways supports epigenetic involvement in obesity-related metabolic stability and energy dysregulation[50].

Our findings also underscore the significant epigenetic impact on lipid metabolism, as evidenced by altered expression of miR-339-5p, miR-328-3p, and miR-29c-3p. These miRNAs regulate lipid uptake, cholesterol synthesis, and macrophage function, potentially through epigenetic modulation of genes involved in lipid storage and mitochondrial function[20,31,51–54].

Enrichment analyses confirmed the involvement of these miRNAs in lipid handling and metabolic homeostasis, further supporting their roles in obesity-driven lipid dysregulation[55–57].

Immune-related pathways were also invoked through the dysregulation of miR-142-5p, miR-1249-3p, and miR-27a-3p. These miRNAs influence cytokine secretion, inflammatory responses, and immune cell activation[58–60], potentially contributing to obesity-induced chronic inflammation through epigenetic modulation of immune signaling pathways, such as Rap1 signaling[61,62].

Additionally, our results highlight the potential influence of obesity-driven EV-miRNAs on neuronal processes, neuroinflammation, and BBB integrity, reflecting the brain-fat axis as a key conceptual framework. Other relevant miRNAs, including miR-29a-3p, miR-142-5p, miR-15a-3p, let-7a-5p, miR-197-3p, miR-296-5p, and miR-550a-3p, were associated with the regulation of neuronal function, synaptic plasticity, and neuroimmune interactions [52,63–67]. Enriched pathways, such as axon guidance, calcium signaling, and glutamatergic synapse regulation, further indicate epigenetic impacts on neuronal communication and cognitive function, potentially exacerbating obesity-associated neurodegenerative risk[68,69].

Glucagon-like peptide-1 (GLP-1) plays a critical role in obesity and is associated with metabolic dysfunction by enhancing insulin secretion, improving glucose metabolism, and regulating energy balance[70,71]. GLP-1 agonists have come into sharp focus as promising anti-obesity drugs in adults[72]. However, only a limited number of studies have investigated their therapeutic potential in pediatric obesity, highlighting a critical area for further clinical exploration[73,74].

Intriguingly, several dysregulated miRNAs identified in our study, such as miR-29a-3p, miR-29c-3p, miR-142-5p, miR-27a-3p, miR-15a-3p, and let-7a-5p, have established regulatory roles in GLP-1 signaling pathways. Specifically, the miR-29 family (including miR-29a-3p and miR-29c-3p) modulates insulin secretion and β-cell function, essential components of GLP-1–mediated glucose regulation[75–77]. MiR-27a-3p directly targets and negatively regulates the GLP-1 receptor (GLP1R), thereby impairing GLP-1 signaling, reducing insulin secretion, and worsening glucose homeostasis[78]. MiR-142-5p, known for its role in inflammation and β-cell dysfunction, indirectly impacts GLP-1–mediated signaling pathways[79]. MiR-15a-3p is implicated in insulin secretion and has been proposed as a predictive biomarker for response to GLP-1 receptor agonists, underscoring its clinical relevance in metabolic control [79–81]. Furthermore, let-7a-5p directly binds to and suppresses GLP1R expression, consequently impairing GLP-1 responsiveness, insulin sensitivity, and glucose metabolism mechanisms closely linked to metabolic disturbances observed in obesity[3]. Given these established interactions, the dysregulation of these miRNAs in pediatric obesity suggests potential impairment in GLP-1 signaling, providing novel epigenetic insights and potential therapeutic targets for enhancing GLP-1 responsiveness in obese pediatric populations.

The clinical urgency of addressing pediatric obesity is underscored by recent global projections indicating that obesity among children and adolescents is expected to continue rising, with an anticipated 360 million individuals aged 5–24 years affected by 2050[3]. Importantly, the forecasted prevalence spike, particularly in younger children (aged 5–14 years), accentuates the urgency of this global crisis. This increase highlights not only the necessity for early detection and targeted interventions but also potential long-term societal and economic consequences. Most alarming is the reality that, despite marked advances in our understanding of the pathobiology of obesity and the era of GLP-1 agonists in mitigating adult obesity at the individual patient level, all regions of the world have failed to curb these disturbing pediatric obesity trends[3]. Current approaches are failing a generation of children and adolescents[3]; therefore, novel and minimally invasive biomarkers, such as circulating EV-miRNAs, are urgently needed.

Collectively, our findings provide intriguing insights into the theranostic potential of EV-miRNA epigenetic regulation of metabolic homeostasis, immune function, and neuroinflammatory pathways, thereby implicating these nanoparticles as critical mediators within the brain-fat axis.

## Clinical Relevance and Future Directions

Our study highlights circulating EV-miRNAs as promising minimally invasive biomarkers for pediatric obesity, enabling early detection, risk stratification, and personalized monitoring of therapeutic responses. The stability and accessibility of EV-miRNAs in plasma underscore their clinical relevance for real-time assessment of metabolic and inflammatory status in pediatric populations.

These investigative field opens new avenues for miRNA-targeted therapeutic interventions, whereby modulation of specific dysregulated miRNAs might help restore metabolic, immune, and neuronal homeostasis in obesity. Given the substantial economic and societal impacts forecasted due to the growing pediatric obesity burden globally, translating these EV-miRNA signatures into clinical biomarkers and targeted therapies represents an urgent and high-impact opportunity for public health interventions. Preclinical validation through animal models and mechanistic studies will be essential to substantiate the therapeutic potential of these miRNAs. Longitudinal studies examining dynamic EV-miRNA responses to dietary, pharmacological, or surgical interventions could further solidify their clinical applicability as dynamic biomarkers and therapeutic targets.

Future research involving larger, diverse pediatric cohorts, coupled with functional epigenetic analyses, will be critical for validating these miRNA-mediated mechanisms and translating these discoveries into personalized clinical strategies. Integrating EV-based epigenetic biomarkers into clinical practice may significantly enhance precision medicine approaches, ultimately improving outcomes for pediatric obesity and its associated metabolic and neuroimmune complications.

## Limitations

This study has several limitations. Firstly, the small sample size may limit statistical power and generalizability, necessitating validation in larger cohorts. Secondly, the cross-sectional design restricts causal inference and assessment of longitudinal EV-miRNA dynamics. Additionally, although matched by age, sex, and BMI, we did not control for other potential confounders such as pubertal stage, diet, physical activity, medication use, and socioeconomic status, which could independently affect circulating EV-miRNA expression [82]. Lastly, the computational predictions of miRNA targets and pathways require experimental validation and additional mechanistic studies to confirm the pathobiological relevance and clinical applicability of EV-omics.

Addressing these limitations in future studies will enhance the clinical applicability and therapeutic potential of circulating EV-miRNAs as biomarkers and targets for pediatric obesity-related metabolic and neuroimmune dysfunction.

## Methods

### Ethics Approval

This study involves human participants and was approved by the University of Virginia (UVA) Institutional Review Board for Health Sciences Research (study ref ID: 20877, date: 08/14/2018). All participants provided informed consent prior to the start of any study-related procedures.

### Study Participants

Pediatric subjects were enrolled into a prospective study conducted at the University of Virginia. Participants were classified as obese (pOB, n = 3) or healthy-weight (pNW, n = 3) and matched by age and sex. Group BMI stratification followed CDC percentiles for healthy weight (BMIs ≤ 85th%tile) and overweight/obese (BMIs >85th%tile).

### Thrombin treatment for plasma defibrination

Plasma samples from participants underwent thrombin treatment to eliminate fibrinogen, thereby facilitating subsequent EV isolation. Specifically, 250 µL of plasma was incubated with 2 µL of thrombin (550 U/mL, Cat# 605157, EMD Millipore, Burlington, MA, USA), mixed gently, and allowed to react at room temperature for 5 minutes. The fibrin clot was then pelleted by centrifugation at 10,000 × g for 5 minutes, and the clear, defibrinated supernatant was carefully collected without disturbing the clot.

### EV isolation via Tangential Flow Filtration (TFF)

EVs were isolated from defibrinated plasma samples using a robust TFF approach. Plasma samples were initially passed through 0.2 µm polyethersulfone (PES) syringe filters (MilliporeSigma, Burlington, MA, USA) to remove residual cellular debris. The filtered plasma was concentrated to approximately 5 mL and adjusted to a final 7 mL volume with sterile phosphate-buffered saline (PBS, pH 7.4) to maintain consistency and prevent EV loss. TFF utilized a custom-built system with a 500 kDa molecular weight cutoff (MWCO) hollow fiber filter (Repligen, Waltham, MA, USA). Plasma circulation through the system was maintained at 35 mL/min via a peristaltic pump (Cole-Parmer, Vernon Hills, IL, USA). Diafiltration involved continuous PBS addition, effectively eliminating low-molecular-weight contaminants while preserving EV integrity. Post-TFF, EV concentrates were further purified using 30 kDa MWCO centrifugal filtration units (MilliporeSigma Amicon Ultra; Fisher Scientific, Pittsburgh, PA, USA) via centrifugation at 4000 × g for 30 min at 4°C. Final EV samples were resuspended in 100 µL sterile PBS for downstream analyses.

### Microfluidic Resistive Pulse Sensing (MRPS)

Particle size distribution and EV concentration were determined using MRPS technology (Spectradyne nCS1, Torrance, CA, USA) with C-400 cartridges, capable of detecting particles within 65–400 nm diameter ranges. Data analyses were conducted using nCS1 Data Analyzer software.

### Transmission Electron Microscopy (TEM)

Plasma-derived EV ultrastructure was examined by TEM. Formvar/Carbon-coated copper grids (200 mesh, Cat #FCF200-Cu; Electron Microscopy Sciences, Hatfield, PA, USA) were plasma-treated before EV application. Grids were incubated with 10 µL EV sample for 1 min; excess liquid was blotted, and grids underwent sequential washing steps in distilled water. Negative staining was performed with UranyLess contrast stain (Cat #22409; Electron Microscopy Sciences) for 22 seconds, followed by blotting and overnight drying. Imaging utilized a Tecnai TF-20 electron microscope (FEI, Hillsboro, OR, USA) operated at 200 kV.

### Western blotting

EV samples were lysed in radioimmunoprecipitation assay (RIPA) buffer (Thermo Scientific, Waltham, MA, USA) supplemented with protease and phosphatase inhibitor cocktails (Thermo Scientific, Waltham, MA, USA) and incubated on ice for 15 minutes. Protein concentrations were quantified using the Micro BCA™ Protein Assay Kit (Thermo Scientific, Waltham, MA, USA). Equal protein amounts were mixed with Laemmli sample buffer containing 2-mercaptoethanol (Sigma-Aldrich, St. Louis, MO, USA), briefly heated, and separated via sodium dodecyl sulfate-polyacrylamide gel electrophoresis (SDS-PAGE) on 4–20% Mini-PROTEAN® TGX Stain-Free gels (Bio-Rad, Hercules, CA, USA). Proteins were transferred to polyvinylidene fluoride (PVDF) membranes (Bio-Rad, Hercules, CA, USA). Membranes were blocked and incubated overnight at 4°C with primary antibodies against CD63, CD9 (Abcam,Cambridge, MA, USA) and Calnexin (Novus Biologicals, Centennial, CO, USA), diluted in Tris-buffered saline containing 0.1% Tween-20 (TBS-T). After thorough washing, membranes were incubated for 1 hour at room temperature with horseradish peroxidase (HRP)-conjugated secondary antibodies. Protein bands were visualized using Clarity Max™ Western ECL Substrate (Bio-Rad, Hercules, CA, USA), and images were captured using the Bio-Rad ChemiDoc™ MP Imaging System.

### RNA extraction, miRNA sequencing and bioinformatics

RNA isolation, miRNA sequencing, and bioinformatics were conducted by Creative Biostructures (Shirley, NY, USA).

Total RNA was extracted from EV samples using Trizol reagent (Invitrogen, CA, USA). RNA quality (RIN >7.0) and quantity were verified using Bioanalyzer 2100 and NanoDrop spectrophotometry (Thermo Fisher Scientific, MA, USA). Small RNA libraries were generated with the TruSeq Small RNA Sample Prep Kit (Illumina, San Diego, CA, USA). Libraries underwent PAGE-based size selection, quantification, and quality control prior to single-end sequencing (50 bp) on an Illumina HiSeq 2500 platform.

### Bioinformatics Analysis

The raw sequencing data underwent comprehensive processing using a customized bioinformatics pipeline. Initial preprocessing involved trimming adapter sequences, discarding low-quality reads, and removing sequences of common RNA contaminants, including ribosomal RNA (rRNA), transfer RNA (tRNA), small nuclear RNA (snRNA), small nucleolar RNA (snoRNA), as well as repetitive and low-complexity sequences. Subsequent analyses focused on unique sequences ranging from 18 to 26 nucleotides, aligning these reads against precursor sequences cataloged in miRBase 22.0 using BLAST to classify known and novel miRNAs. Alignments permitted slight length variation at both the 3’ and 5’ ends, with allowance for a single mismatch.

Sequences aligning with mature miRNA regions within the hairpin arms were classified as known miRNAs. Those mapping to the opposite arm of known miRNA precursors, but not previously annotated, were designated as novel 3p- or 5p-derived miRNAs. To explore potential homologous miRNAs, reads that did not map initially were further aligned against other precursor miRNA sequences in miRBase 22.0, followed by genomic alignment to determine exact locations within the reference genome. Remaining unmatched sequences underwent additional genomic alignment, and potential miRNA hairpin structures were identified using RNAfold software (http://rna.tbi.univie.ac.at/cgi-bin/RNAWebSuite/RNAfold.cgi).

Stringent criteria guided secondary structure predictions, including constraints such as maximum bulge size in stem regions (≤12 nucleotides), minimum stem region base pairs (≥16), free energy threshold (≤ -15 kcal/mol), overall hairpin length (≥50 nucleotides), and loop length (≤20 nucleotides). Further refinement included restrictions on mature miRNA regions such as maximum nucleotides in a bulge (≤8), maximum biased errors (≤4), limited biased bulges (≤2), overall error allowance (≤7), and a minimum base-pair requirement within mature regions (≥12). Additionally, mature sequences were required to occupy at least 80% of the predicted stem region.

### Differential expression analysis

Differential expression analysis of miRNAs was conducted to identify significantly upregulated and downregulated miRNAs between PC versus pMS. Differences were compared using t-test with BH adjustment for multiple comparisons. Given our small sample size, we chose a less stringent p-value ≤ 0.1 to define statistically significant miRNAs but also evaluated differences with p-values of 0.001, 0.01 and 0.05. FDR-adjusted q-values were also obtained.

### Volcano plot analysis

Differential miRNA expression analysis identified significantly upregulated and downregulated miRNAs. Results were visualized using volcano plots, with log2 fold change (log2FC) plotted against the negative logarithm of the p-value (-log10 p-value). Significantly altered miRNAs were identified by applying thresholds of |log2FC| ≥ 1 and p-value ≤ 0.05, highlighted distinctly in red for upregulated and blue for downregulated miRNAs, whereas non-significant miRNAs appeared in gray.

### Scatter plot analysis

To assess the correlation and distribution of miRNA expression levels between groups, scatter plots were generated. Normalized expression values from paired conditions were plotted against each other, with each point representing an individual miRNA. A trendline provided insight into the overall correlation pattern, helping identify consistent expression trends and outliers across the datasets.

### Venn diagram analysis

Venn diagrams were employed to illustrate overlaps and unique miRNAs among different experimental groups. This approach enabled visualization of shared and condition-specific miRNAs, facilitating the identification of commonly regulated miRNAs as well as those uniquely modulated under specific experimental conditions.

### Pathway analysis

To further elucidate biological roles and regulatory functions of the differentially expressed miRNAs, pathway enrichment analysis was conducted using the Kyoto Encyclopedia of Genes and Genomes (KEGG) and Gene Ontology (GO) databases. Predicted miRNA target genes were identified through public databases such as TargetScan, miRDB, and miRanda. Statistically enriched pathways were visualized using bar charts and network diagrams, providing comprehensive insights into relevant biological processes, molecular functions, and cellular components potentially influenced by the identified miRNAs.

## Data availability

All data generated and analyzed in this study are included within this manuscript and the supplementary information file. Additional data requests may be directed to the corresponding author.

## Author contributions

Conceptualization: MDH, ER, JNB, SMM

Study design and funding acquisition: ER, JNB, SMM

Investigation and conducted research: MDH, JNB

Organization and data analysis: MDH, SMM

Completed the original draft of the manuscript: MDH, SMM

Data interpretation, revision, and manuscript correction: MDH, ER, JNB, SMM

All authors approved the final version of the manuscript

## Additional information

Correspondence and requests for materials should be addressed to Senior Corresponding author.

## Acknowledgements

SMM was supported by intramural funding from the Abigail Wexner Research Institute at Nationwide Children’s Hospital and NINDS (5K12NS098482-05). ER was supported by National Institutes of Health (NIH) grants UG3/UH3TR002884 and U18TR003807. JNB was supported by NINDS (K23 NS116225). The content is solely the responsibility of the authors and does not necessarily represent the official views of the National Institutes of Health.

