## Supplementary information for "Circulating extracellular vesicle-miRNAs as epigenetic mediators of metabolic, inflammatory, and neurological pathways in pediatric obesity"

700 Children’s Drive

Columbus, OH 43205, USA.

***Co-corresponding Author:** J. Nicholas Brenton, MD

Address:	University of Virginia

Division of Neurology, Division of Pediatric Neurology,

PO Box 800394

Charlottesville, VA 22908, USA.

**Supplementary Table 1. Differentially expressed EV-miRNAs in pNW vs. pOB**

| Downregulated DEMs | FC | log₂(FC) | *P-value* |
| --- | --- | --- | --- |
| miR-643 | 0.1 | -3.27 | 3.20E-03 |
| miR-590-5p | 0.3 | -1.75 | 5.67E-02 |
| miR-103a-2-5p_R+1 | 0.35 | -1.51 | 5.86E-02 |
| miR-542-3p | 0.27 | -1.89 | 6.44E-02 |
| miR-5189-3p | 0.24 | -2.07 | 6.65E-02 |
| miR-142-5p_L+2R-2 | 0.62 | -0.69 | 2.20E-02 |
| miR-27a-3p_R-1 | 0.57 | -0.8 | 4.62E-02 |
| miR-330-5p_R-2 | 0.68 | -0.56 | 7.21E-02 |
| miR-24-3p_R-2 | 0.67 | -0.58 | 7.22E-02 |
| miR-618 | 0.13 | -2.95 | 7.51E-02 |
| miR-6866-5p | 0.23 | -2.1 | 7.58E-02 |
| miR-27b-5p_R+1 | 0.55 | -0.87 | 8.08E-02 |
| miR-548ag_R+1_1ss4GA | 0.57 | -0.8 | 8.09E-02 |
| miR-561-5p | 0.39 | -1.36 | 8.10E-02 |
| miR-4645-3p_L+2R-1 | 0.28 | -1.85 | 8.33E-02 |
| miR-361-3p | 0.4 | -1.32 | 8.81E-02 |
| miR-4772-3p | 0.58 | -0.79 | 9.01E-02 |
| miR-210-3p | 0.51 | -0.97 | 9.04E-02 |
| miR-6813-5p | 0.38 | -1.41 | 9.05E-02 |
| miR-223-5p_R+2 | 0.59 | -0.76 | 9.23E-02 |
| mir-1302-1-p5_1ss9AG | 0.17 | -2.58 | 9.44E-02 |
| miR-605-3p | 0.61 | -0.72 | 9.74E-02 |
| Upregulated DEMs |  |  |  |
| miR-550a-3p | 34.2 | 5.1 | 4.86E-02 |
| miR-1273c_R-1 | 5.31 | 2.41 | 5.11E-02 |
| miR-1268a | 6.36 | 2.67 | 3.43E-02 |
| miR-1268b_R-2 | 6.36 | 2.67 | 3.43E-02 |
| miR-1249-3p | 4.43 | 2.15 | 8.67E-03 |
| miR-3173-5p | 3.29 | 1.72 | 4.56E-02 |
| miR-296-5p | 3.23 | 1.69 | 3.08E-02 |
| miR-328-3p | 3.11 | 1.64 | 1.76E-02 |
| miR-15a-3p_1ss22AT | 3.1 | 1.63 | 3.22E-02 |
| miR-5010-3p_R+1 | 2.93 | 1.55 | 1.49E-03 |
| miR-197-3p | 1.91 | 0.93 | 3.05E-02 |
| miR-339-5p | 1.56 | 0.64 | 2.15E-02 |
| let-7a-5p | 1.54 | 0.63 | 4.69E-02 |
| miR-454-3p_R+1 | 1.74 | 0.8 | 5.45E-02 |
| miR-106b-5p | 1.79 | 0.84 | 5.71E-02 |
| miR-15b-5p | 1.49 | 0.57 | 6.17E-02 |
| miR-30b-5p | 1.47 | 0.55 | 6.92E-02 |
| miR-766-3p | 2.94 | 1.56 | 6.95E-02 |
| miR-224-3p_L-2R+1 | 3.56 | 1.83 | 7.04E-02 |
| miR-1296-5p | 1.26 | 0.33 | 7.10E-02 |
| miR-342-3p | 2.83 | 1.5 | 7.58E-02 |
| let-7g-5p | 1.61 | 0.69 | 7.61E-02 |
| miR-744-5p | 1.36 | 0.44 | 7.62E-02 |
| miR-671-3p | 1.56 | 0.64 | 7.70E-02 |
| miR-20a-3p_R+1 | 2.54 | 1.35 | 7.72E-02 |
| miR-548k | 1.62 | 0.69 | 8.72E-02 |
| miR-15b-3p_R-1 | 1.67 | 0.74 | 9.07E-02 |

DEM, Differentially expressed miRNAs; FC, fold change; pediatric healthy control normal-weight; pNw, pediatric obese pOB
